# Rapid antigen testing for SARS-CoV-2 infection in a university setting in Ireland: learning from a 6-week pilot study

**DOI:** 10.1101/2021.08.05.21261660

**Authors:** Gerald Barry, Catherine McCarney, Marc Farrelly, Rory Breathnach, Carmel Mooney, Simon J. More

## Abstract

**Objectives:** With the ongoing circulation of SARS-CoV-2 in countries across the world it is essential to identify effective ways to reduce the risk of infection while allowing society to function as close to ‘normal’ as possible. Serial testing using rapid lateral flow antigen tests is a possible way to do this by screening populations in a targeted way, identifying infectious (both symptomatic and asymptomatic) people and removing them from circulation while infectious. To make rapid antigen testing effective, high levels of participation are important. This study was designed to evaluate the establishment of a testing programme in a university setting and assess some of the factors that impact participation in such a study among both staff and students.

**Study Design:** Observational, survey

**Methods:** A trial period of SARS-CoV-2 rapid testing using the Abbott Panbio rapid antigen test was set up and staff and students based in the University College Dublin Veterinary Hospital were asked to take part voluntarily for 6 weeks. Following the trial period, we used a questionnaire to evaluate satisfaction and to understand some reasons behind participation or lack thereof.

**Results:** Overall, almost all respondents to the survey stated that they were happy with having a testing programme present in the workplace and it helped to reduce anxiety associated with COVID-19. Findings indicated that staff and students did not participate equally in the voluntary testing programme. The findings also highlighted that intrinsic motivations and extrinsic motivations for participation differ. For example, participation among staff was much higher than among students, motivational messaging focused on protecting others did not resonate with students as much as staff, convenience was a key factor driving participation in both cohorts and the pressure of being forced to miss class (if positive) close to exam time provided motivation to students to avoid testing.

**Conclusions:** Introducing antigen testing into a workplace helped to reduce overall anxiety associated with the potential impact of COVID-19, but achieving good participation was challenging. Participation is key to a successful, campus wide antigen testing programme but reaching high levels of participation is not straightforward and can not be taken for granted. Different motivations drive participation in different cohorts and different messaging/incentivisation is needed to encourage participation in those different cohorts. The findings reported here should inform any SARS-CoV-2 testing programme that will run in these types of settings in the future.

## 1. Introduction

The COVID-19 pandemic that started in China in late 2019 has caused disruption to the whole world in one form or another. The betacoronavirus, SARS-CoV-2, that causes COVID-19, primarily transmits between people in aerosol droplets, so close contact in poorly ventilated indoor environments are areas of high risk of transmission if an infected person is present. In Ireland, as elsewhere, the isolation of infectious individuals is central to efforts to limit SARS-CoV-2 transmission. People with symptoms are asked to self-isolate, however, this does not remove the risk posed by pre-symptomatic and asymptomatic individuals. Pre-symptomatic transmission is known to be substantial ^1^ and asymptomatic or mildly symptomatic individuals may also shed virus at levels sufficient to infect others ^2^. There is considerable heterogeneity in estimates of relatively infectiousness of symptomatic and asymptomatic people, and no conclusive evidence of difference ^3^. Further, SARS-CoV-2 does not shed from infected people equally, with approximately 2% of infected people carrying approximately 90% of the total virus load in a population at any one time. The distribution of viral loads, by PCR test, cannot be distinguished according to symptoms suggesting that people shedding large amounts of virus are as likely to be asymptomatic as symptomatic ^4^. Viral load is associated with increased likelihood to shed infectious virus, so those people with the most virus in their upper respiratory tract are the most likely to infect others and to be associated with superspreading events ^5^. Collectively, in the context of effective disease control, these factors highlight the need to identify those with the highest virus loads, regardless of their clinical presentation.

Antigen-detecting rapid diagnostic tests (Ag-RDTs) offer a means to rapidly identify infected individuals, with the potential to substantially reduce the risk of onward SARS-CoV-2 transmission. However, like every diagnostic test, rapid antigen tests are not perfect and there are both strengths and weaknesses with them. An Ag-RDTs using a lateral flow device (LFD) has a lower sensitivity than PCR. In a detailed review, sensitivity of Ag-RDT was estimated to be 72.0% (95% confidence interval 63.7-79.0) and 58.1% (40.2-74.1) for symptomatic and asymptomatic individuals ^6^, respectively, but noting that sensitivity does vary between test manufacturers ^7^ and will be operator-dependent ^8^. Although Ag-RDTs have a higher risk of false negatives that PCR, infected people test positive by LFD during a relatively narrow window of the infectious period, coincident with the period of highest viral load (see figure 1 ^8^) when individuals are likely most infectious ^9^. Ag-RDT results are informative at a single point in time, and will not, for example, test positive for those in their latent period (0-3 days following initial infection approximately). On the other hand, most Ag-RDTs have very high specificity, recently estimated at 99.5% (98.5-99.8) and 98.9% (93.6-99.8) in symptomatic and asymptomatic individuals respectively ^6^.

**Figure 1.**
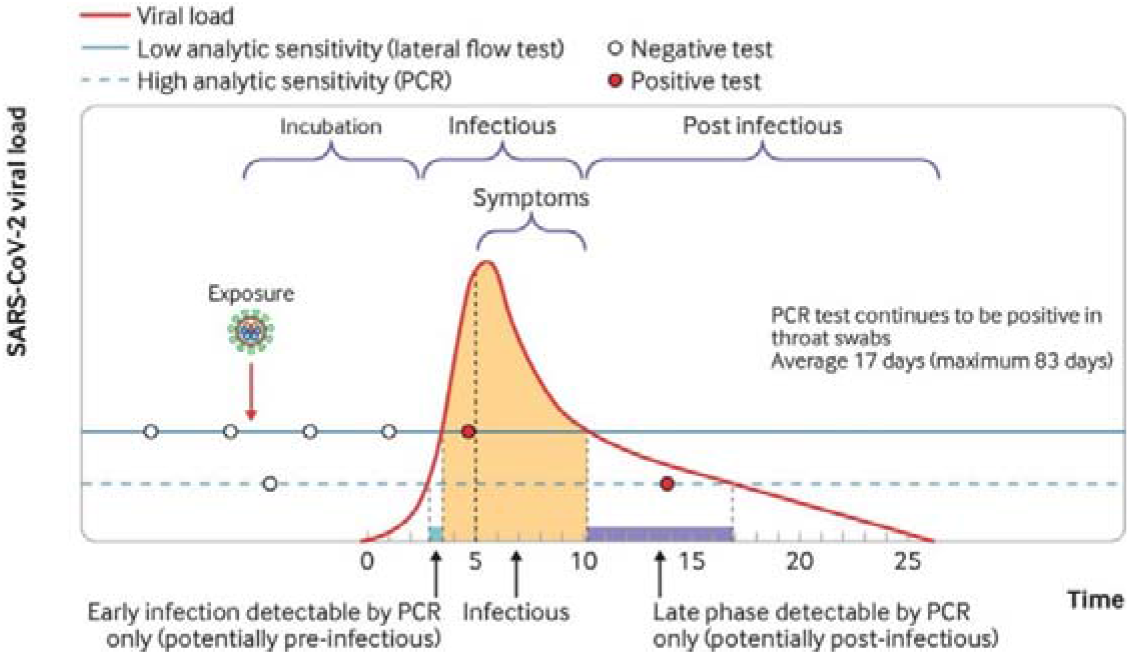
An illustration of the average time taken to reach peak viral load and its association with symptoms and the sensitivity of both the PCR test and the antigen test. Taken from ^8^

**Figure 1.**
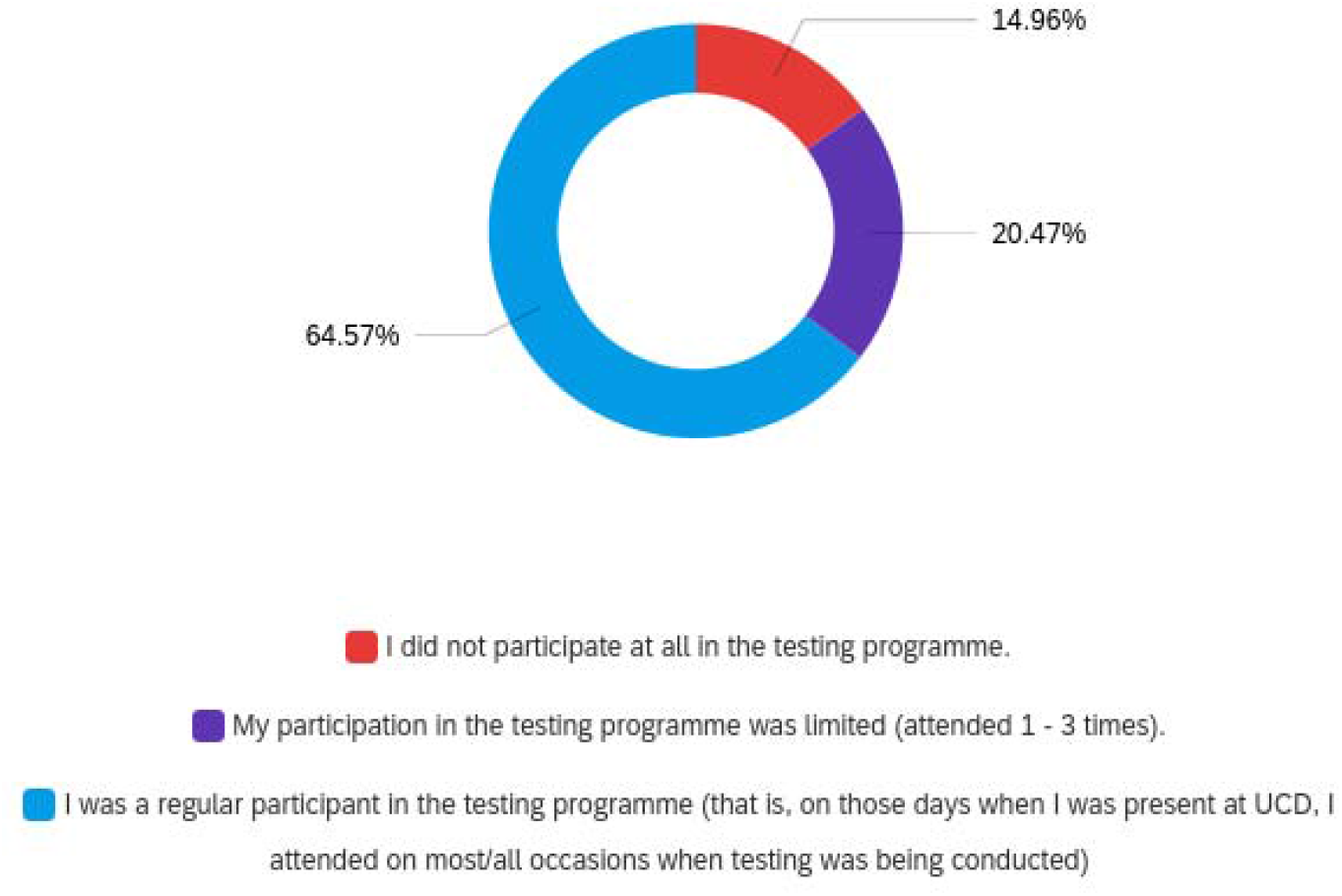
Answers to the question: ‘Which of the following best describes your level of participation in the COVID-19 rapid antigen testing pilot?’ 127 people responded.

The World Health Organisation recommends the use of Ag-RDTs in specific situations, including outbreak investigation/contact tracing, monitoring trends in disease incidence, widespread community transmission and to test asymptomatic contacts of cases (World Health Organisation, 2020). In contrast, the use of Ag-RDTs can be problematic in populations with low infection prevalence and low pre-test probability. In these settings, the positive predictive values will be low (leading to a high proportion of false positive results) when testing is conducted at large-scale ^7^. In situations where the pre-test probability is low-to-moderate, Ag-RDT results must be interpreted with care. In these settings, a Ag-RDT positive should be considered preliminary, with individuals isolating until a PCR test result is known. Ag-RDT negative individuals should continue to comply with conventional risk mitigation measures ^10^. Ag-RDTs will be most effective when used regularly, with high levels of participation.

In the UK, Ag-RDTs have been used in universities, schools and care homes ^11,12^ to rapidly identify – and remove – those asymptomatic people who are shedding high amounts of virus. Early results from a pilot at the University of Liverpool were disappointing, with approximately half of cases missed (a sensitivity in comparison to the PCR of 48.9%, specificity of 99.93%) ^11^. In Ireland, there is now considerable experience in the use of Ag-RDTs to detect SARS-CoV-2 infection in meat processing plant workers, which is recognised as a high-risk setting. In these situations, once or twice-weekly Ag-RDTs has been suggested as a viable alternative to once-monthly RT-PCR serial testing, specifically with the aim to identify and isolate asymptomatic individuals ^13^.

This study was conducted to investigate the feasibility and limitations of an antigen testing programme conducted in a university environment which was considered at higher risk of SARS-CoV-2 transmission.

## 2. Methods

### a. The setting

University College Dublin (UCD) Veterinary Hospital is a critical teaching resource for the UCD School of Veterinary Medicine staffed with approx. 120 clinicians and postgraduate students. It also accommodates a series of clinical rotations for approximately 170 final year veterinary and veterinary nursing students for much of the academic year. The veterinary hospital remained open throughout the pandemic, handling almost 10,000 cases in 2020, but was considered at higher risk of SARS-CoV-2 introduction and spread, in part due to the density of staff and students and the nature of the work (in terms of close contact with staff and animal owners) when handling and treating animals appropriately. A pod-based approach was implemented in early 2020 but could not be sustained once hospital caseload returned towards normal levels, in large part because of broader health and safety concerns associated with limited staff numbers. From March 2020, a range of mitigation measures were put in place following detailed risk assessment, including a review of student rotations, PPE, physical distancing etc. From 2021, further measures were considered, given the serious national picture (7-day incidence, levels of community transmission), the emergence of SARS-CoV-2 variants, and in light of staff anxieties in the context of returning student rotations. As a consequence, and in the interests of business continuity, there was further evaluation of final year teaching, changes to teaching timelines, the introduction of higher specification face masks and the strategic placement of air purifying units. Further, as reported here, a pilot study of testing using a Ag-RDT was conducted with veterinary hospital staff and students over a 6-week period from 24 March to 29 April 2021, coinciding with a national 14-day incidence of confirmed COVID-19 cases of between 159.54 and 127.26 per 100,000 population at the start and the end of this period, respectively.

### b. Recruitment of participants

An information drive was established prior to the start of the pilot study, including information videos, circulation of written information and a webinar to explain the principles of the test and the reasons for doing the pilot. Veterinary hospital staff and students received an initial invitation and follow-up reminders during the first three weeks of the study. Participation was voluntary, and all participants were required to read and sign a consent form (supplementary file 1) before their first test. The study was conducted with approval from the UCD Human Research Ethics Committee, approval number LS-21-20-Barry.

### c. The testing regime

The Panbio™ COVID-19 Ag rapid test device (Abbott, Ireland) was used for all tests. The nasal test swab was self-administered under supervision from a trained tester. Training videos and were also provided in advance of the first day of testing. Swabs were then tested using a lateral flow device by a trained tester independent of the person that had been swabbed. All test swabs were labelled with a code so testers did not know who they were testing.

Testing was carried out each Monday and Thursday. A person being tested entered a designated room, they were directed to a swabbing station, and they were then instructed how to perform the swab. The swabbing was then observed to ensure correct technique before the swab was placed in a test tube for analysis by the testing team. The swabber being tested was allowed to leave after being swabbed and their swab was analysed within 30 minutes of the swab taking place. If the person was negative, they were not contacted, but if they were positive, they were informed verbally as soon as the result came through. A rubric for how a positive person should act after testing positive was generated and used to guide candidates that tested positive.

### d. Participation

For staff, audit was conducted on Monday and Thursday of week four of the study to determine the total number present in the hospital and eligible to participate in testing. Staff participation was calculated for each of these two days. For students, rotation information was reviewed to determine the number present at each of 8 days during weeks 2-5 of the study. Average student participation over these 8 days was determined.

### e. Follow-up questionnaire

At the end of the 6-week trial, a link to an electronic survey (supplementary file 2) was sent by email to all staff and students that could have taken part in the study. Importantly, this included both those that participated and those that did not but were eligible to do so.

## 3. Results

### a. Participants

A total of 159 people attended at least once for testing during the pilot, including 112 staff and 47 students. The staff were working in a variety of roles within the veterinary hospital, whereas the students were all final year veterinary medicine or veterinary nursing students.

### b. Test results

A total of 798 tests were performed across a 6-week period with no positives recorded. No member of staff or of the student cohort reported any symptoms associated with COVID-19 or tested positive by PCR independent of the trial. Staff participation was 90% and 75% on the 2 audited days, whereas the average student participation was 19%.

### c. Survey results

A total of 128 people participated in the survey, including 88 staff and 38 students.

#### Level of participation

When asked about their level of participation, the 127 people who responded to this question said they had participated as often as possible, however, 20% participated less than 3 times, and 15% said they had not participated at all (Figure 1).

#### Reasons for limited or no participation

Of the 19 people who said they didn’t participate, 13 provided reasons for non-participation, and of the 26 people who participated between 1 and 3 times, 21 provided reasons why they did not participate more. Collectively, the reasons for limited or no participation among these 34 respondents included the following: 1 said they were not interested, 1 said they didn’t want to risk a positive result, 8 said they did not have enough time, 2 people chose not to attend because they felt they were at low risk and didn’t need to attend, and others indicated that their time on campus was limited [11] or when on campus they did not have time to participate [7 people]. The remainder [4 people] said that they simply forgot to attend, even when on campus.

#### Opportunities to increase participation

There were a total of 81 respondents to this question, and answers can be grouped into the following 4 themes:

- **Reminders:** Many people indicated that a reminder on the day of the test would have been useful. This could have come as a text message or an email, but forgetting to attend was clearly an issue for many.
- **Convenience**: It was mentioned by a number of people that making the swabbing more convenient would have allowed better participation. Allowing self-swabbing at home and dropping the swab upon arrival at work was brought up on a few occasions. Having the swabbing station at the front door of the hospital or within the hospital itself so people are reminded to do it and not inconvenienced was also suggested.
- **Time:** This was commonly raised as an issue. Solutions to this focussed on broader testing windows – running testing earlier or perhaps later (after 5pm) so people did not have to take time out of work. It was also suggested that a dedicated break could be introduced so that people could go to the swabbing station without feeling like they were missing work. It was also suggested by some students that they were reluctant to leave their post if no other staff were going, so a dedicated testing time, or timings outside of work hours would have worked better.
- **Incentives:** It was mentioned on a few occasions that students were reluctant to attend because of the fear of missing 2 weeks if they tested positive. Despite emphasis on keeping fellow workmates safer by getting tested, this was not incentive enough for many. With this in mind, a number of respondents pointed out that incentives might encourage more participation. Incentives such as the ability to use a testing certificate to be allowed to do more things, partake in more activities. Other incentives such as sweets, a bottle of wine for the most participation etc. were also mentioned.

#### Further comments

A total of 58 people responded to this question. Answers were provided as free-text, but can be summarised as follows:

- Reduced anxiety among workers.
- Created a feeling of an extra layer of safety in the workplace and worked well alongside all other mitigations such as mask wearing, hand sanitisation and increased ventilation.
- An understanding that it reduced but didn’t remove the possibility of a superspreading event in the workplace.
- Great initiative.
- Friendly and welcoming.
- Good information and training.

These comments are summarised in the word cloud generated from the answers (figure 3)

**Figure 3.**
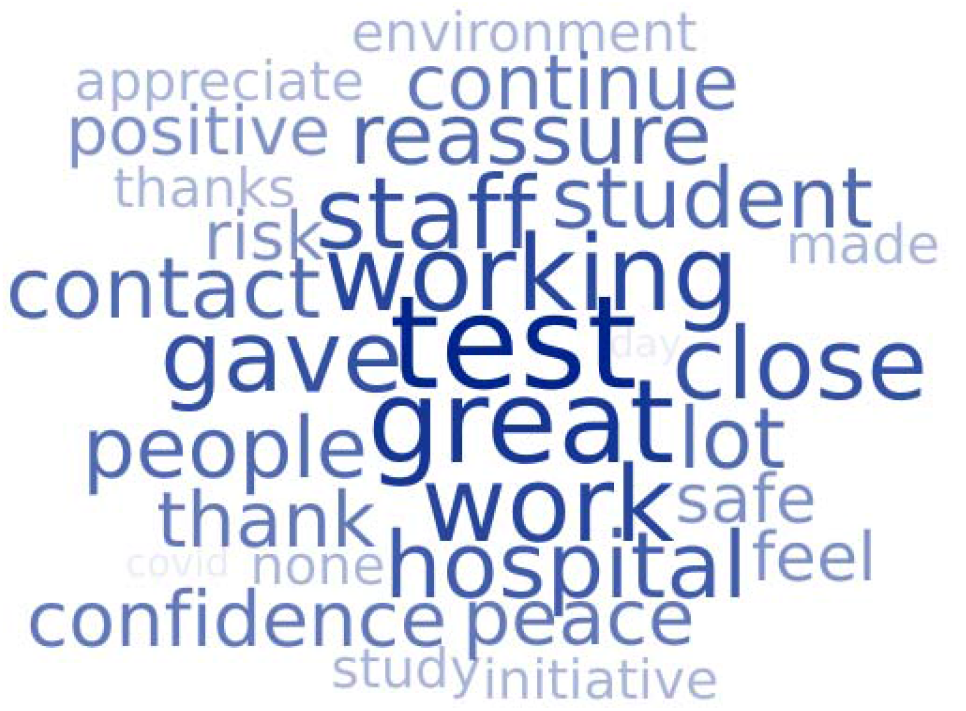
Word cloud generated from the answers to the question asking respondents to leave any other comments they might have about the antigen testing pilot.

#### Support for ongoing testing

In total, 98% of the 112 people who responded to this question said they wanted it to continue as long as COVID-19 was a concern (see figure 4).

**Figure 4.**
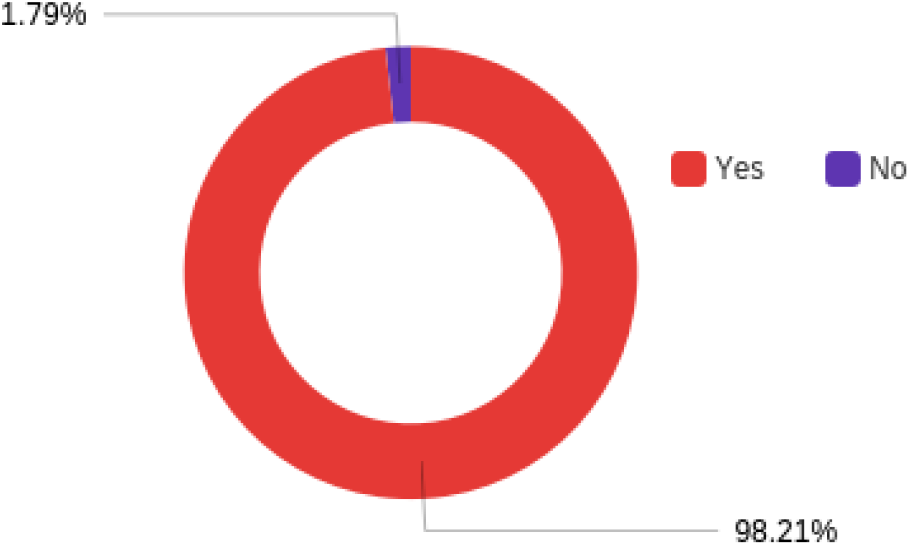
Answers to the question: ‘Would you like to see antigen testing continue until the risk of COVID-19 has decreased substantially?’ There were 112 respondents to this question.

## 4. Discussion

### a. Summary

Antigen testing has a role to play in reducing the overall risk of infection in a population but it is important, as with any diagnostic test, to ensure that the testing is carried out correctly and effectively. The current study assessed the feasibility and limitations of an antigen testing programme conducted in a university environment which was considered at higher risk of SARS-CoV-2 transmission. The testing programme was conducted on a regular basis over a 6-week pilot with a relatively large group of workers and students.

### b. Feasibility, including lessons learned

The pilot study was implemented successfully, and a relatively large group of staff and students were tested on a regular basis in a safe and efficient manner over the 6-week period. Further, as a result of the trial, the participants reported a decrease in anxiety in the workplace and increased confidence among workers. The testing programme allowed participants to carry out their work aware that an extra layer of risk reduction was in place alongside other risk mitigations such as high-grade masks, physical distancing, hand cleanliness and increased ventilation. Importantly, the introduction of the antigen testing was not associated with a noticeable decrease in people following HSE guidelines around the other risk mitigations, so risk of a COVID-19 outbreak in the workplace was decreased overall.

There was excellent feedback from staff and students in relation to the information they received beforehand as well as the training videos. Nonetheless, the initial swabbing technique among participants varied greatly, highlighting the importance of on-hand assistance for first-time participants. Correct nasal swabbing technique will likely improve test sensitivity and it is vital that this aspect is performed correctly every time. The current study found that after some initial one-to-one direction, technique became relatively uniform and efficient.

The strengths and weaknesses of rapid antigen tests were clearly highlighted to participants, including prudent interpretation of test results. In particular, an understanding that while a positive means that the individual is very likely to be infected and shedding virus, a negative test means that virus was not detected at this point but it did not mean you aren’t infected, and therefore one should continue to follow all other public health guidelines. The major benefit of an antigen test is that it will pick up highly infectious people efficiently and conveniently, but with the possibility of missing infected people. This was clearly explained to participants in the information leaflet. These messages have been informed by earlier experience of rapid antigen testing in Ireland, during ongoing screening of asymptomatic people in meat processing plants in January 2021 ^13^. Over 5000 people were tested by both PCR and antigen test, with no evidence of false positive results (a specificity of 100%). In the meat processing plant study, the sensitivity was found to be approximately 80% in people with a PCR Ct below 25 %, falling below 60% in people with Ct values above 30.

In the current study, there were no changes to existing mitigation practices within the veterinary hospital. The overwhelming sentiment from staff and students was that antigen testing provided extra reassurance, and greater confidence that the working environment was a safer place to be. From a mental health and team morale point of view, this was reported as being a successful and important consequence of the study.

### c. Limitations, including lessons learned

High levels of participation are vital to make any testing regime effective. Reliance on people staying away from the University campus when symptoms appear is not sufficient to prevent COVID-19 outbreaks, given that asymptomatic and mildly symptomatic people can also shed virus. Serial testing by either rapid antigen test or PCR is conducted independent of symptoms, focusing instead on the detection of highly infectious people. In order to carry out a test on a serial basis, however, it should ideally be easy to perform, convenient and cheap. It should be cost-effective, and people must be willing to submit to testing regularly.

In the staff cohort, participation levels were high (75-90 %) and commentary about the test was generally very positive with regular use of words such as peace of mind, confidence and safe in the survey. No concerns were raised about the test itself. In contrast, the participation in the student group was very low (19%), which appears to be due to a number of contributing factors. Among students, a common reason given in the survey and also verbally during the study, when queried, was the fear of testing positive and the consequent impact on their studies. The timing of the pilot study may have played a role in this anxiety, as it was the final few weeks of term before graduation, with exams approaching and rotations in the hospital pending that needed completion. Nonetheless, it is worrying that some people would place these concerns ahead of the benefit that would accrue from a positive test result, including the reduction in infection risk, which could potentially be devastating, for fellow staff and students. It would be informative to conduct a similar pilot study at a different time of year, when similar pressures are not present (the start of the academic term for example), to see if this issue dissipates. Another common reported issue was time. Although this was a factor for both staff and students, students may have felt extra pressure, either consciously or otherwise, to avoid missing time in hospital for the sake of a test. It is apparent, based on feedback from both staff and students, that convenience is a priority and would help participants to participate more regularly, particularly when time is at a premium.

Throughout the pilot study, messaging in support of participation had a strong public good element, seeking to reduce infection risk both to the individual and their fellow classmates / workmates / hospital as a whole. While this apparently worked well with staff, it did not appear to resonate with the vast majority of students at our institution. The interplay between public good and private benefit has been considered in several recent papers, with differing results. In a recent study from the US, Thunström et al. (2021) found that healthy younger people were more likely to take a no-cost COVID-19 test than health older people, with people generally selfless in their decision to test for COVID-19 ^14^. In contrast, Fallucchi et al. (2021) found that willingness-to-test was increased in association with altruism, conformism and risk-aversion and decreased with decreasing age and increased willingness to take risks ^15^. Messaging tailored to our students with an emphasis on personal benefits or rewards may have been more successful. Indeed, a bottle of wine was suggested by one survey respondent or sweets for every swab by another. Incentives may be a way to encourage participation in the student cohort and this is something that needs to be considered going forward, although this would then have to be made available for staff too.

### d. Conclusions

In conclusion, the trial showed that antigen testing could be carried out effectively and efficiently in a university setting with a relatively large cohort, on a regular basis. The study also identified differences in participation in the staff and student cohort, suggesting different approaches are needed to incentivise different cohorts. Future studies will focus on promoting how a testing regime can bring personal benefits to students, rather than emphasising the benefit of increasing safety to others. Future work will also focus on convenience and the potential, for example, of running at home testing and self-reporting rather than attending a testing centre.

Because of the concern around participation, it could also be considered to deploy rapid testing during times of case number surges only, rather than on a serial basis during times when case numbers were stable or low. One could initially train large cohorts to carry out self-testing during times when cases were low and if the need arose, because of a surge of cases in a concentrated geographical area, test sites could be rapidly set up to test large numbers of people in a short space of time. While not ideal theoretically, because regular testing would be better as a risk mitigation strategy, practically, surge testing might be more feasible and cost effective.

## Data Availability

All data associated with the study is available upon request.

## Acknowledgements

We would like to thank University College Dublin for supporting this study. We thank all participants, both students and staff of University College Dublin for taking part.

