## Supplementary File 1 for "Rapid antigen testing for SARS-CoV-2 infection in a university setting in Ireland: learning from a 6-week pilot study"

**Consent form for COVID-19 antigen testing**

Yes	No

| I agree to take part in this antigen testing pilot study and understand I can withdraw at any time. |
| --- |
| I have read and understood the information about this antigen test. The information has been fully explained to me and I have been able to ask questions, all of which have been answered to my satisfaction. |
| I agree to swab my nose as instructed and to submit my nasal swab for testing using the Abbott Panbio COVID-19 Rapid Antigen test. |
| I agree that if I am informed that the test detected SARS-CoV-2 antigen then I will immediately go home and phone my GP / student health service (as applicable) to arrange a COVID-19 PCR test. |
| I understand that a negative result does not preclude me from following all COVID-19 related guidance. On occasion, the test can miss a positive case (produce a false negative). |
| I understand that to take part in this testing I agree to swab my nose and submit a sample twice weekly for a 6-week period. |
| I give permission for anonymised material/data to be stored for possible future research **related** to the current study **without further consent being required,** but only if the research is approved by a Research Ethics Committee. |

Signature:

Print Name:
