## Supplementary File 2 for "Rapid antigen testing for SARS-CoV-2 infection in a university setting in Ireland: learning from a 6-week pilot study"

Supplementary file: Complete survey results

**Q1 - Are you a**

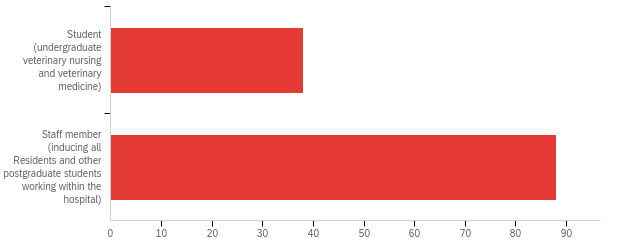

| # | Field | Minimum | Maximum | Mean | Std Deviation | Variance | Count |
| --- | --- | --- | --- | --- | --- | --- | --- |
| 1 | Are you a | 1.00 | 2.00 | 1.70 | 0.46 | 0.21 | 126 |

| # | Answer | % | Count |
| --- | --- | --- | --- |
| 1 | Student (undergraduate veterinary nursing and veterinary medicine) | 30.16% | 38 |
| 2 | Staff member (inducing all Residents and other postgraduate students working within the hospital) | 69.84% | 88 |
|  | Total | 100% | 126 |

**Q2 - How did you originally hear about the hospital COVID-19 rapid antigen testing pilot?**

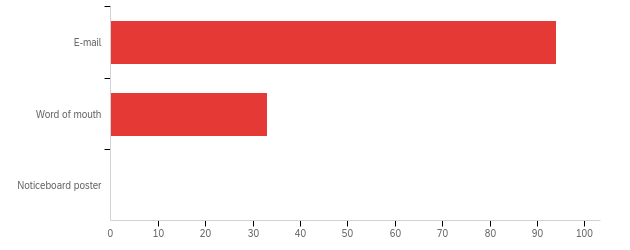

| # | Field | Minimum | Maximum | Mean | Std Deviation | Variance | Count |
| --- | --- | --- | --- | --- | --- | --- | --- |
| 1 | How did you originally hear about the hospital COVID-19 rapid antigen testing pilot? | 1.00 | 2.00 | 1.26 | 0.44 | 0.19 | 127 |

| # | Answer | % | Count |
| --- | --- | --- | --- |
| 1 | E-mail | 74.02% | 94 |
| 2 | Word of mouth | 25.98% | 33 |
| 3 | Noticeboard poster | 0.00% | 0 |
|  | Total | 100% | 127 |

**Q3 - Which of the following best describes your level of participation in the COVID-19 rapid antigen testing pilot**

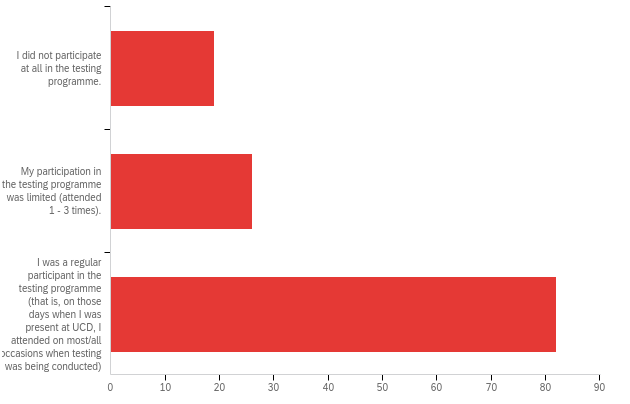

| # | Field | Minimum | Maximum | Mean | Std Deviation | Variance | Count |
| --- | --- | --- | --- | --- | --- | --- | --- |
| 1 | Which of the following best describes your level of participation in the COVID-19 rapid antigen testing pilot | 1.00 | 3.00 | 2.50 | 0.74 | 0.55 | 127 |

| # | Answer | % | Count |
| --- | --- | --- | --- |
| 1 | I did not participate at all in the testing programme. | 14.96% | 19 |
| 2 | My participation in the testing programme was limited (attended 1 - 3 times). | 20.47% | 26 |
| 3 | I was a regular participant in the testing programme (that is, on those days when I was present at UCD, I attended on most/all occasions when testing was being conducted) | 64.57% | 82 |
|  | Total | 100% | 127 |

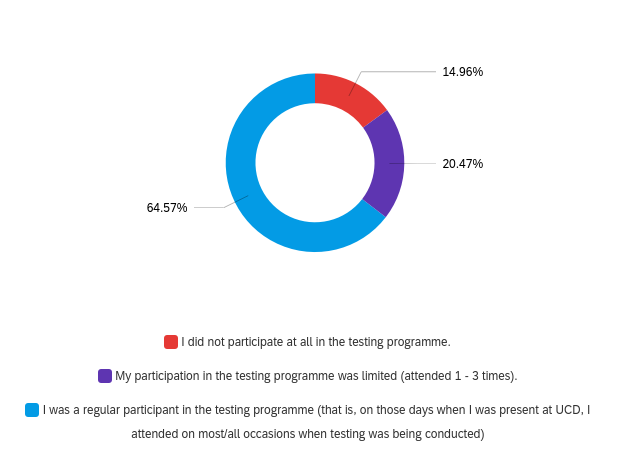

**Q4 - Why did you not participate?**

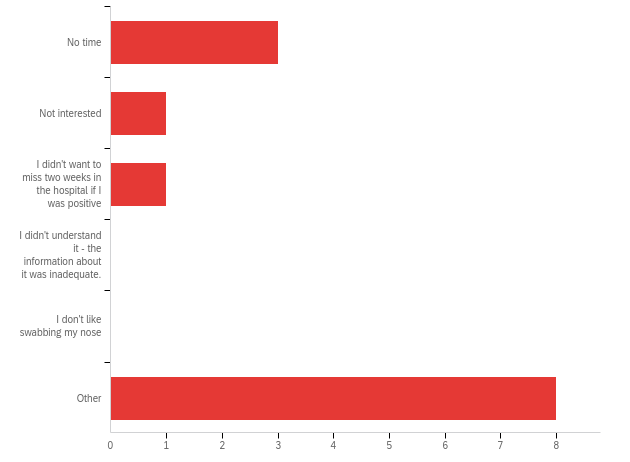

| # | Field | Minimum | Maximum | Mean | Std Deviation | Variance | Count |
| --- | --- | --- | --- | --- | --- | --- | --- |
| 1 | Why did you not participate? - Selected Choice | 1.00 | 6.00 | 4.31 | 2.20 | 4.83 | 13 |

| # | Answer | % | Count |
| --- | --- | --- | --- |
| 1 | No time | 23.08% | 3 |
| 2 | Not interested | 7.69% | 1 |
| 3 | I didn't want to miss two weeks in the hospital if I was positive | 7.69% | 1 |
| 4 | I didn't understand it - the information about it was inadequate. | 0.00% | 0 |
| 5 | I don't like swabbing my nose | 0.00% | 0 |
| 6 | Other | 61.54% | 8 |
|  | Total | 100% | 13 |

Q4_6_TEXT - Other

Other - Text

When the programme started I had two weeks left in the college and I didn’t think I would be the right candidate for the pilot study as it was a 6 week study - I hope that rapid antigen testing is brought out to schools and workplaces as it a good vice to have

I had only FACS left, I felt there was a low risk of transmission between people outdoors and wearing masks.

Finished rotations by the time it was offered so was not around hospital to partake.

I was not present on campus while the trial was being done.

I wasn't on rotation at the time

I had left Ireland

Usually was not available for testing at the times it was being performed during rotations. When I was off rotations I didn't want to make 2 trips on public transport to come to UCD for testing as I wanted to limit my exposure to other people.

Already had Covid-19

**Q5 - What could have been done to encourage you or make it easier for you to participate in the testing programme?**

What could have been done to encourage you or make it easier for you to participate in the testing programme?

If it was a case scheduled in the day where everyone had to attend as I wasn’t entirely sure it’s only in the final week I saw geralds email saying everyone to join and I will take part if and when this is rolled out nationally

Time assigned on rotations to go and get swabbed. I was not in the hospital for very long while the testing was being carried out as most of my rotations had been completed by then but the time I was in the hospital was quite busy and I did not have an opportunity and at times simply forgot to go during the testing time.

Mandatory testing

Offer an antibody test for previous exposure in return for participation in the antigen test, even at a small cost would drum up interest to get the ball rolling.

Definitely would have taken part if I was in the hospital at the time of conducting testing.

Had I been on campus I would have willingly participated

If I had been on rotation I would have definitely participated

Make it « valuable » eg certificate as virus-free that may be used at airport etc

Provide set time for it during rotations

The timings weren't suitable when we were on rotations. I did not want to miss out on already reduced learning time to participate. Testing before the start of a rotation would have been easier.

Start it earlier

More available times, or scheduled tests during rotations so that we aren't asking to leave during procedures/rounds etc.

Wasn't sure if it would show up as a false positive which can happen after recovering from coronavirus

nothing

**Q6 - Why was your participation was limited?**

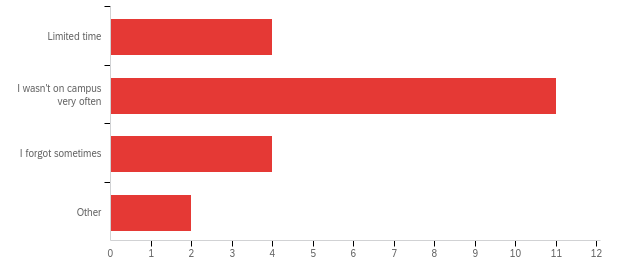

| # | Field | Minimum | Maximum | Mean | Std Deviation | Variance | Count |
| --- | --- | --- | --- | --- | --- | --- | --- |
| 1 | Why was your participation was limited? - Selected Choice | 1.00 | 4.00 | 2.19 | 0.85 | 0.73 | 21 |

| # | Answer | % | Count |
| --- | --- | --- | --- |
| 1 | Limited time | 19.05% | 4 |
| 2 | I wasn't on campus very often | 52.38% | 11 |
| 3 | I forgot sometimes | 19.05% | 4 |
| 4 | Other | 9.52% | 2 |
|  | Total | 100% | 21 |

Q6_4_TEXT - Other

Other - Text

Didn't get any notification of testing times after the first 3 weeks

Night shifts and hollidays

**Q7 - What could have been done to encourage you or to make it easier for you to participate more in the testing programme?**

What could have been done to encourage you or to make it easier for you to participate more in the testing programme?

It was very easy to participate not sure if much more could have been done .

I found the weekly reminder emails really useful and a good reminder.

If it would have been possible, the ability to take swabs home and do them when suits, then drop the sample in to UCD

Have potentially more days or another testing window i.e. afternoon slots. Sometimes my schedule wouldn't permit making the timeframe

I think most of the non participation from vet students came from worries of what would happen should they test positive

Greater range of times for testing. Ideally even an hour after end of regular work day (eg 5-6pm) would have been better than current availability

Send out the swabs to students?

The programme started just as I was finishing my rotations. I was only in for one week and I went for the test during this one week on campus

See above

Possibly afternoon hours as well

more visibility in school and hospital of testing

nothing I just needed to take the habit but every one was very nice and arranging and the procedure is veruy quick

Later appointments-afternoon slots

Nothing, I just was off clinics. I think the people did a good reminder Every Antigen testing day.

I don't think there was anything difficult with the organisation, it just need to get into the habit of it.

will partake when time permits

send SMS

Calendar reminder or text message

**Q8 - What aspects of the testing programme worked, well in your opinion?**

What aspects of the testing programme worked, well in your opinion?

All

Easy to be carried out

the programme was well thought out and organised

The walk in service, the location in the school

Flexibility in attendance times through the session

The quickness of it

Time slots were very good, the whole process was very efficient and quick

The open access times - morning/lunch time fitted best with clinics. Easy place to find, friendly staff.

Rapid

Hours, organisation

Very quick and easy

It was very quick very well organised and made me feel much safer at work during COVID times

It was a very speedy process, so i wasnt away from work for long.

It was really quick, really easy to do yourself and the timing was mostly very flexible

Good instruction videos provided beforehand.

Very organized and socially distanced! Excellent instructions given - very easy to do.

Speed of the process

Fast safely carried out

It was quick and easy to perform. Easily accessible in the vet building. I liked how after the first 2 weeks there were no scheduled individual time slots. While on rotation, I had missed 1 or 2 tests initially because something came up in the hospital e.g. a patient's anesthesia recovery went very long...etc. This way I could go get my test when I was able.

Organised and efficient

I think it was fast and easy

Very quick and easy to do. I could get in and out in 5 mins

Flexible timing

Very efficient

The open times (rather than booked) for the hospital life much better, and made me able to come more

Having set time slots allowing students to work it into our day and alert any supervisors of our need to leave

Quick and easy

The nurse in charge of the rotatin we were on (almost) frogmarching us upstairs to make sure we had the test (the unit was v busy at the time but she stopped all students doing anything until we'd had our test!

Very time efficient, well spaced out and no waiting around there was never more than 2-3 people ahead of me.

Flexibility with time slots

All, I thought it was well run.

The efficency of the team and convenience of it not only being in the early morning as I start at a later time

Ease of participation

organisation, speed of testing

The fact it was a walk in service with a wide time range

The walk in format, self swabbing, fast and easily accessible

Walk in service a hard to get off the floor for a set time

well run and quick to do

quick - nice personel - efficent

The speed at which testing could be done

Quick

Regular testing

Free schedules

Easy to access, no waiting around, process was quick and painless

Testing was simple and quick, I appreciated knowing whether I was covid negative

Very well organised

Easy, no waiting

efficient , safe, with current numbers I dont thin booking appointment necessary and hard to keep time when on clincs

The whole testing programme worked extremely well and gave me great peace of mind. At first booking the time slots was a bit difficult but the walk in allocated times worked very well.

Fast and efficient

Easy to perform the test, rapid and very close to the hospital building

The time range when it was available

The easy and flexible access for testing was perfect, especially regarding the variable work day in the hospital

Knowing we are working safer

Continuty and how easy and speedy it was

Walk in clinic was very helpful, no waiting time and fast process

Very well organised; quick, no wait

The ease of access. Drop-in at anytime.

drop in clinic - helpful staff

no appointments just walk in no waiting

The instruction videos and the accessible system

I thought the whole programme worked very well

Not having to book in just drop in at certain times worked very well

Room allocation and space

Consistently, simplicity and the fact it was actually available

Very quick and well organized

location,

Very well organised

Walk-in availability - helps when on clinics and unable to guarantee when you will have a free couple of minutes

The testing system was fast

Quick and easy

It was quick and efficient. Provide more confidence we are in a safe environment

Well organized. Quick. Compatible with the busy clinical day almost all the days

Straightforward procedure, good guidance from staff running testing

Very quick and efficient process

Really easy access, very quick, never had to wait or queue. Very simple process that took a couple of minutes max really.

Very quick only took a couple of minutes

How quick it was to do.

The ease of doing the test

Extremely quick process

Separation, easy access

Everything went very smoothly

Easy to follow steps and quick test to do

Ease of testing and quick procedure.

I found it worked very well

Ease of access

It’s simple and very quick

all aspects - very efficient, very helpful, very friendly

The test itself is easy to perform and was quick.

Flexible hour, rapid to perform. Very easy.

test was quick, easy and non invasive. Didnt take much time away from work day and rotation system in and out worked really well in my eyes as there where never people lining up nor was there multiple people in the room.

All

Twice weekly testing

Streamlined, with access early in the morning before clinical work began

**Q9 - What aspects of the testing programme could have been improved, in your opinion?**

What aspects of the testing programme could have been improved, in your opinion?

None

limited hours on days if testing. perhaps consider less stations and longer hours for testing

n/a

Use names instead of numbers

Just hard to remember to come to the test when busy with work

Making it mandatory

Weekly emails/reminder would have been great

Nothing - I thought it went very smoothly.

None

Time slots, generally the hospital is busier in the mornings so it can be difficult to get to the morning slot and perhaps the midday slot with an afternoon one wouldve been better

More regular testing

More awareness and reninders

n/a

None

Na

Not sure..

Reminders of days and times via email the morning of

More advertising on days of testing it is easy to forget when you’re in the hospital

Wider range of times available, or more days for testing

Nothing really, it was very simple and everyone was friendly

If possible having a wider testing period because often things happening in the hospital would prevent me going to an original slot and trying to reschedule for a different day

started much earlier during the covid pandemic - eg Sept 2020

Increased/alternative hours, nursing student night shifts are 7am-7pm so a later time slot that would allow them to get it done before they start would be useful as I spent 2 full weeks unable to partake. I wouldn't have minded arriving early e. 5:30-6pm if it meant I could participate

Pre booking wasn't convenient

Honestly can't think of any.

I wouldn't say it needs improvement but I don't work the full week so only got tested once a week. I am very glad to have had to get the chance to have a test once a week but I still would have been in work a few days without getting a test done

Often I was out on visits in the mornings so that is the only reason the timings didn't work for me

An option for an earlier test would be good as could not attend if in surgery for example.

Maybe a slightly earlier start at 8 am

Overall I thought was very well ran, increased time slots would be beneficial

Email communication did not seem to reach all staff members

testing area in the hospital closer to workplace

none

Longer opening hours for testing for busy days

More reminder emails for walk in tests

More Testing

None

Afternoon appointment slots. I would often be busy in the morning or in later and when I rememberd it was usually after lunchtime.

Sometimes I forgot to attend, regular reminders are useful

None

Maybe more reminders in the hospital for staff

N/a

For me the testing programme worked perfectly. I would also be very happy to self test if this was a possibility

Timing. It was more difficult to attend the early morning slots.

Maybe just an email reminder every Monday with time and days of testing

I can't thinck of anything. It worked very flawless. I liked the reminder emails (once the email wasnt send and I directly forgot)d

More participation particularly from students

Not sure

none

None

it was well done

Maybe there is scope to 'graduate' the participants to do the test themselves and then drop off the labelled tube at a designated point (eg outside the clinpath lab on Tuesday and Thursday)./ This would eliminate the requirement of people to be present

None

It all ran smoothly

I felt it worked really well

Clearly I missed some times as not on site but not realistic to expect a 24/7 service

Was all good

longer opening times as mornings are busiest times

Available times to get tested.

I always forgot the days on which testing was happenning

Later slots, hard to get away when its busy

None

None

None

location was difficult to when on clinics. I wish it were closer to the hospital

If possible maybe the first testing slot could be a little earlier. Then maybe students could be encouraged to go up before they start with cases in the hospital or before rounds.

Nothing- maybe more days for testing, ie more of the same so could access other than Monday or Thursday if couldn't make those days.

Longer notice given if closed early or opening later etc

It worked better when there were no appointment times as it was easier to attend. The break during the morning was a slight inconvenience

It worked very well

Longer testing hours

N/A

information - e.g. I wasn't aware that the testing was still conducted this week and therefore missed Monday.

Extended afternoon times - difficult during busy periods to leave work

n/a

Mosr reminders ...on a busy day its very east to forget

If possible - extended times.

Time of testing staff really busy first thing in the morning and consults etc going on at second time

to be give a sweet at the end

Sometimes I forgot about it until later in the morning because there was no e-mail reminder and the testing is done off-site, as it were

Make it more mandatory to increase attendance and significance of the results.

Nothing that i can think of.

N/A

Nothing, it was great

**Q10 - We are always looking to increase participation - what in your opinion could have been done to improve attendance among staff or students?**

We are always looking to increase participation - what in your opinion could have been done to improve attendance among staff or students?

More reminders & communication to remind people of the importance

not sure. Perhaps emphasised that results would not be disclosed apart from participant that tested positive

n/a

Reminders in the hospital (notices etc) as during a busy day it was easy to forget you had to go

Email reminders on each test day were great & the only way I remembered to go. If i missed the email i missed the test

Keep sending the emails and spreading the word

Incentive. Bottle of wine for the person with the most tests. Seminar for staff on benefits of regular testing?

Nothing

as above, mornings are busy, generally when I attended the morning slot it was when I was on a later shift so I went before starting

I did forget to attend one or two sessions so a morning reminder on the day of testing woukd be ideal

Text reminders would work effectively

More regular email notifications might help

Variety of times e.g. afternoon session? Although times from week3 onwards suited me well.

If rotations supervisors or clinicians encouraged students to step out of their rotations briefly to go get tested that would be great. I.e. if they said "ok the testing window is open, anybody who wants to go is free to go and come back"

Longer testing hours or closer to vet hospital

Maybe some kind of reminder? e.g. text or email alert on the day of testing

Above

More assurance to people as I think not getting tested came from a place of worrying what would happen if you tested positive

For people I talked to the issue was mostly with the time of year (large number of students travelling at the end of rotations, heavy rotation schedule, preparation for exams)

Encouraged more by clinicians

A lot of students were saying “I don’t want to get tested, what if I test positive?!” And that made no sense to me, because wouldn’t you rather test positive and have the chance to protect people than be positive AND put more people at risk? Like you’re positive either way, so I think maybe something needs to be done to bring that home because the fear of testing positive and having your schedule disrupted should not outweigh the fear of BEING POSITIVE and CAUSING HARM. Address the scheduling fear by offering support, and reassurance, and remind them that peoples lives are more important than schedules

More information on the running of the actual test as well as having wider testing windows

Free lollipops

I'm not aware what the participation was so I can't answer this question

Students didn't want to sign up because they thought they'd have to travel in on weeks they weren't on rotations

Notifying people during rounds etc the days testing were occurring or reminder emails that morning

Members of staff in the hospital encouraging students to go.

We have had a few very busy weeks in work so I suppose it is hard to even get upstairs to get the test done. I wonder if it was set up in the hospital like h038, would more people go in and you would see people going in and that may remind you

Again just timings

I think several students were unwilling to attend because they felt they had no clarity regarding what would happen if they tested positive - specifically that they did not know if they would have to repeat a rotation, which would have resulted in delayed graduation. A clear message from the School would be necessary to make clear they would not be disadvantaged by a positive test. For staff, I think most attended, but some mornings, they were not available.

To email on Wednesday evening as well to remind for the Thursday slot.

Increased number of time slots. If testing was available half an hour earlier I think increased numbers of residents and interns could participate prior to starting work. There were a number of mornings where staff intended on getting tested were waiting for testing to open but then became caught up in procedures in the hospital.

more testing areas

Reminder messages on testing days

Reduce work load would give more time to participate

Better Knowledge of Testing

None

Text reminder option?

Regular reminders

More days available during the same week

More info on why the test is beneficial

hospital can be very busy at times and I know my colleagues wanted to attend but making way upstairs limited them- I think a testing center downstairs would have made it easier for them to attend

Perhaps email reminders or posters on the doors as a reminder as on some days I only remembered as others were going.

Email notifications

See previous answer

I think it was overall organised well

I got the impression the students werent aware they are able to particapate from the beginning. Maybe communicate this even more clear.

Cover being provided more easily

Longer hours due to crazy case loads in the hospital

Regular alert reminders, text reminder. When working in the hospital during busy times it was easy to forget and miss the clinic times

More frequent reminders

Reassurance for students on clinical rotation regarding rearrangement protocols if positive test. Staff already high participation

Reminder emails in the mornings when tests were conducted

By the tiem the testing started on the 25th of March the students had been on rotations since the fist of Feb without any major outbreaks so I think they were less worried and therefore the uptake was lower. The timing in relation to the working hours of staff improved after the first week but still would catch out those starting at 7.30 and by 9.30 already being involved with intakes and procedures. More 'independent' testing like above would help those.

Regular reminders

More days with drop in testing

Increase in Staff/ student emails on testing days and if possible increased opening hours

If I could test at home and submit sample when I came in

longer opening times

Longer testing times available as the hospital can get very busy and the most available time where people could go to get tested was when it wasn’t available. Maybe setting up a testing station closer to the hospital floor

Maybe a visible remainder somewhere in the building?

Testing am and pm

Posters

Make it manditory

More reminder emails

I think students were a bit reluctant as were concerned if they tested positive they would miss some days of rotation and with the limited rotations this year they would have had no time to make up days before exams. I don't know if it could be made a mandatory health and safety component of rotations - similar to how it is mandatory to wear appropriate PPE in the hospital.

I really wish it could be mandatory but understand the medico-legal hurdles here. I'd hope that these could be ironed out before extending the project. I had to repeatedly remind students to avail of it, not sure why they didn't uptake it more easily. EG, in a group of 14 only 4 of them signed up so had to bug them by email to emphasize it so I think then the participation improved. Maybe was there an element of being so close to the end of the year they were reluctant if by any chance they tested positive and then couldn't continue to exams? I'm not sure what the reason was but I do think it should be mandatory for F2F participation in rotations. When I talked to students about it they were all enthusiastic and fully accepting of it.

A bigger emphasis on making sure colleagues have the time or get cover to run up and get swabbed

Perhaps utilising a room downstairs so it’s even quicker for hospital staff to be tested

Don't see any incentive necessary. Highlight importance of keeping your and those around you Covid free

Reminder emails on the day of testing

Posters on doors

More word of mouth notice as not everyone reads emails

Instagram/Facebook reminder posts? Posters around school? Weekly emails?..

Times beyond what were available.

Not advertised in hospital enough in hospital some students were not even aware of it

to be give a sweet at the end

E-mail reminders on the day, posters within the hospital, testing on way into hospital

Making it mandatory to enter the building or attend rounds in the hospital.

incentives for people who do it regularly

Give chocolate for attending

I think it was really well done

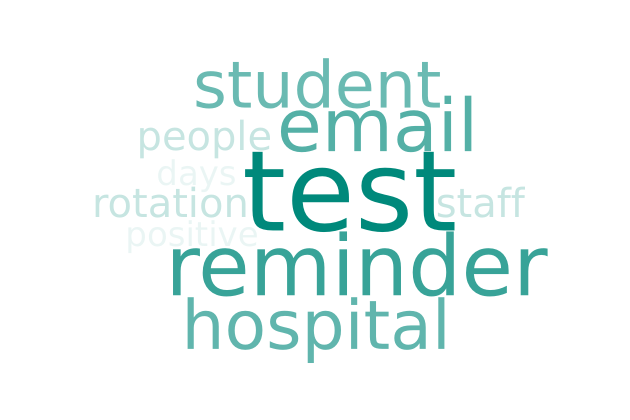

**Q11 - Would you like to see antigen testing continue until the risk of COVID-19 has decreased substantially?**

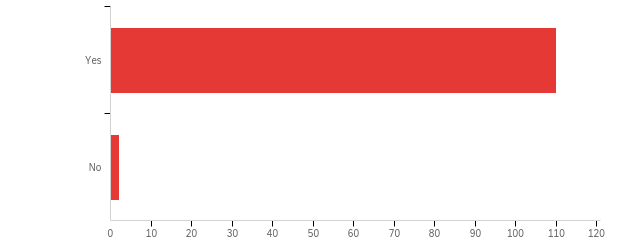

| # | Field | Minimum | Maximum | Mean | Std Deviation | Variance | Count |
| --- | --- | --- | --- | --- | --- | --- | --- |
| 1 | Would you like to see antigen testing continue until the risk of COVID-19 has decreased substantially? | 1.00 | 2.00 | 1.02 | 0.13 | 0.02 | 112 |

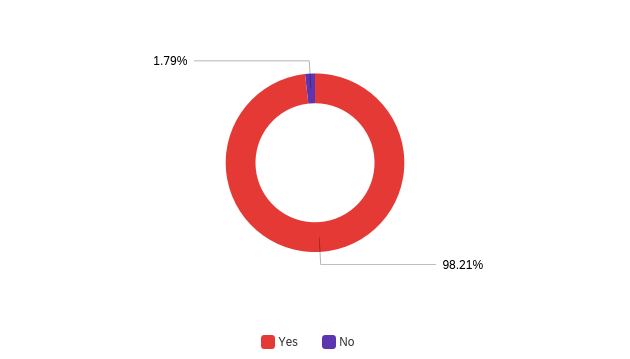

**Q12 - Any other comments about the COVID-19 rapid antigen testing pilot study?**

Any other comments about the COVID-19 rapid antigen testing pilot study?

I reminded many staff members each day the testing was on & it was frustrating that not more staff members took part reliably, especially as some people are better at PPE compliance than others. I felt that having the testing in place gave a sense of reassurance.

I think the testing programme worked very well, and make staff and students feel safer giving that social distancing and other measures are difficult to maintain in close clinical working environment

Great peace of mind personally

Please can we now have the vaccine?

None at the moment

No

I think it would be great to continue particularly because in the hospital we are often in unavoidable close contact it was quite reassuring

I appreciated reassurance my family and i got from taking the test every week

It's an incredibly helpful service to have which reduces the worry from working in such close contact with many people every day

It was a great initiative and helped me to have more confidence that I was safe in the hospital and (hopefully) also not putting others at risk.

n/a

Makes working with so many people in close proximity in hospital less worrying

Thank you, I hope it continues!!

While not as accurate as PCR, it is a helpful and speedy indicator of the presence of microbes

Thank you

How « efficient » / reliable is it, compared to tests run in labs ?

I think it is a fantastic idea and well done to all involved. It made our last few weeks of university a lot less stressful knowing I've had negative tests regularly

really good thing to do - is essential for everyones safety

N/A

No

I greatly appreciate all the work that went into the study so that we could get tested. Thank you

No

This study markedly increased the confidence regarding the safety of work under current circumstances, particularly as we often work in close proximity with different people, including students and staff who variably adhere to guidelines outside of work with respect to social distancing and other measures. It did not lead to any laxity in the approach to PPE, social distancing (when possible) etc at work.

I thought was well run, a lot of positive feedback from hospital staff

it gave me more confidence that a super-spreader of covid would be picked up, reduced anxiety in workplace

Thanks a lot!!

Gave me more peace of mind at work! Great job!

No

Thanks to all who worked on the study

None

Makes working in close contact with ever changing students and staff a lot more reassuring as to risk of covid - despite protocols PODS do not work in the hospital and we are often within 1 foot of each other for over 15 min periods

Thank you to all the testers, I really appreciated getting the testing done and your help!

Great initiative, made everyone feel safer at the hospital.

No, thanks a lot for this brilliant and well executed initiative.

It was great to provide some piece of mind.

Please continue!

Thank you for doing this!

antigen testing is so important, it reassures the staff and students that UCD is doing everything in it's power to provide a safe working and learning environment during the pandemic particularly when the nature of our work prevents us from maintaining physical distancing from others.

This is a great idea and positive in relation to the confidence for staff and students working throughout the pandemic - especially in relation to clinical staff.

it gave some level of reassurance that myself and none in the pod was positive

The testing did reduce anxiety for those staff having a lot of close interaction with both students and other staff, while having vulnerable people at home. With a lot of the staff being tested regularly it gave that bit more security after all of us working in close contact all throughout the pandemic, with PPE, bit still...

Very pleased it went ahead - gave me a peace of mind while working in a busy and close contact environment

Great work- well done all

I genuinely appreciated the fact that testing was made available to me, free of charge and with support from all my colleagues

All people assisting the testing were always very friendly and welcoming.

The opportunity of being tested was fantastic, it gave me the security of coming to work safely and with the peace of mind of knowing that if I was positive I would know. It also gave a great impression of UCD to people that I commented about the testing. My family and friends were very impressed by the timing of testing and the security this gave. It made me feel proud of working in UCD

I believe this was a really positive development which gave a lot of peace of mind to staff members in particular. I really hope it can be continued.

I felt this was a very well run and organised project. It was accessible, quick and involved minimal interruption to the working day. It really helped bolster confidence in the risk mitigation steps put into place and I feel strongly that it should be continued, especially since out teaching environments are so very different to any other teaching course on campus (ie, long days of constant nose to nose contact in wet labs or live surgeries). It was an important element in convincing an external teaching location (DSPCA) to allow students return to live surgery there- without this the learning outcomes of a final year module would never have been attained and students could not have graduated. Thank you very much Dr. Gerald Barry and the team (Marc F, Catherine McC etc) for doing this so well and being so very helpful at all stages of it.

None

I felt it was a proactive approach to managing the risk and feel it could identify incidences of Covid before clusters are allowed develope

My participation was limited to 4 times because I was off clinics most of the time. If the testing continued, I would be very happy to participate.

Brilliant to have on site in the hospital and great for self reassurance

n/a

I think testing should be maintained for the foreseeable future.

I thought it was fantastic idea, simple and gives peace of mind.

good job

This has been a great pilot study and it is hoped that it is viewed positively. Because of the way we work, we cannot avoid close contact, and the antigen testing gave us confidence that together with all other risk mitigation efforts, we were doing all we possibly could to remain open as a Veterinary Hospital.

Great initiative. Thanks for putting it together.

N/A
